# High burden of anemia and malnutrition in two tribal populations of Northeast India

**DOI:** 10.1101/2023.01.27.23285091

**Authors:** Joycy Vungneihchoi, M.P. Sachdeva

**Author notes:** **Corresponding author** (JV). These authors contributed equally to this work.

## Abstract

**Background:** Anemia is a deficiency of red blood cells and a public health burden in India. There needs to be more reporting of the prevalence of this condition in adults consisting of both males and females in a rural setting. Hence, this study is undertaken to address this gap.

**Materials and methods:** 1460 participants were recruited during a household survey in the Churachandpur district of Manipur. Data on personal, social demographic, and lifestyle variables were collected along with anthropometric measurements. Anemia status was tested with the help of a standardized portable hemoglobinometer.

**Results:** The prevalence of anemia was 42% and 46.4% among Kuki and Paite tribal communities of Manipur, respectively, which is significant within the communities. Type of occupation and lifestyle factors were also found to contribute to anemia. Age was also found to be inversely proportional to the prevalence of anemia.

**Conclusion:** The present study found a high prevalence of anemia and malnutrition, a matter of concern. The studied population, the tribals, are the deprived section of society that needs to be taken care of to achieve the United Nations Sustainable Developmental Goals (SDGs). They stay in remote areas which are not easily accessible, and hence they should be prioritized in terms of health and various other developments. Also, this high prevalence of anemia can lead to various health complications like cardiovascular diseases if not treated. Iron supplements should act as an intervention for the high prevalence of anemia and should be delivered timely to vulnerable populations.

## Introduction

According to the World Health Organization (WHO), anemia is a grave worldwide health problem and can be defined as a low hemoglobin concentration. It is a good indicator of reduced nutrient consumption and poor health, which can lead to stunting, wasting, reduced weight at birth, and increased weight during the lifetime as individuals constantly fatigue [1,2]. It is also known to cause mental problems, reduce immunity, and negatively impact the quality of life besides deficit oxygen supply to various body tissues [3]. There are different types of anemia; common causes could be nutrients, iron, folate, vitamin B12 and A, hemoglobinopathies, and diseases such as malaria, HIV, and tuberculosis [4].

Globally, anemia affects 1.76 billion people [5]. The current worldwide prevalence of anemia in 2019 is 29.9% among women of reproductive age, 29.6% in non-pregnant, and 36.5% in pregnant women [6]. The region reported to have the second highest burden of anemia is South Asia, after Western Sub-Saharan Africa [7]. The total burden caused by anemia was calculated in 2019, resulting in 58.6 (40.14-81.1) million years of life with a disability [8]. Although the prevalence of anemia indicates a declining trend in 2019 (22.8%) compared to that in 1990 (27%), different regions still show an increasing trend.

The prevalence of anemia in India as per NFHS-5 (2019-21) was 57% among reproductive-age women, while it is 25% among men, which was higher than that of NFHS-4 reports [9]. In Manipur, NFHS-5 report states that the prevalence of anemia increased for non-pregnant women (26.4% to 29.3%) and decreased for men (9.5% to 6%) when compared to the NFHS-4 report [10]. Malnutrition is an abnormal consumption of nutrients where intake can be deprived or in excess leading to non-communicable disease [11]. Underweight and obese are the types of malnutrition considered in this study. Being Underweight can be defined as a low intake of nutrients, while obesity is the over-accumulation of fats. These abnormalities can act as a risk factor for non-communicable diseases [12]. Globally, 1.9 billion adults are obese, and 462 million are underweight [6]. Such abnormality is a gray side among tribal populations, causing a risk of various cardiovascular diseases [13], and prevalence is high due to their traditional practice and low literacy [14]. In India, the prevalence of underweight men and women, as per NFHS-5 report is 16.2% and 18.7%, respectively, while that of overweight and obese are 22.9% and 24%, respectively [9]. As for Manipur’s prevalence of underweight among men and women (age 15-49 years) is 8% and 7.2%, respectively, showing a decreasing trend compared to the previous NFHS-4 report [10]. There is a rapid increase in the prevalence of overweight men (30.3%) and women (34.1%) compared to the previous report, respectively [10].

According to the 2011 census, the tribal population in India consists of 8.6% of the total population, while in Manipur, 41% of the population belongs to tribals. The word “ Tribal” is self-explanatory because they are deprived of health facilities, education, sanitation, and basic amenities[15]. Although different tribal communities in India are diversified, they still share a common burden of morbidity and mortality [16]. Various studies have been done, yet studies among tribal populations are scarce. Tribals are endogamous populations. Studies of anemia on tribals are vital as they are listed among vulnerable groups, have different cultures and beliefs, and illiteracy and isolation from non-tribal inhabitants expose them to different health and social issues [17]. Several studies on children and women have been done on anemia, yet, there is a research gap in reporting the same for males. Furthermore, most researches deal with urban populations. Hence, the present paper focuses on reporting the prevalence of anemia alongside analyzing the double burden of malnutrition in both males and females of rural settings among two different tribal communities of Manipur.

## Materials and methods

### Study area and people

The study area is a hilly area covered with forest where tribals reside. There are 33 recognized tribal communities of Manipur, and they are broadly divided into two categories – Kuki-Chin-Mizo and Naga. The present study focuses on two tribal communities, Kukis and Paites, that fall under the Kuki-Chin-Mizo group. The Paites in the present study settled in the higher altitude areas of Churachandpur, while the Kukis reside comparatively in the lower altitude region and closer to the heart of the district. The two studied tribal communities mainly depend on jhum cultivation, and rice is their staple food. Yet, a gradual transition in the lifestyle of tribals was observed in terms of occupation, dietary habits, and other lifestyles. Cultivators started shifting to rearing animals and business and took a keen interest in education. Dietary habits were also drastically changed, accompanied by a sedentary lifestyle.

Basic amenities such as water for drinking and household usage are stored in a common water storage tank in each village which is carried in pots to their own houses. Irregular electric supply to rural areas is prominent and poorly affected by the weather. Generally, as per works of literature, the consumption of alcohol and tobacco products is high.

### Study design and recruitment of participants

The present cross-sectional study was conducted among two tribal communities of Manipur. A total of 40 villages were picked randomly, and a house-to-house survey was carried out. Pre-informed written consent was taken from the study participants before recruitment. Ethical clearance was obtained from the Institutional Ethics Committee, Department of Anthropology, University of Delhi. The two tribal communities were selected based on the highest tribe proportion, namely Kuki (24.6%) and Paite (6.6%) from two blocks of Churachandpur district, Manipur.

The sample size was calculated using an online sample size calculator called ‘Sampsize.’ A reported prevalence of 38% [18] was used, an error is 5%, and 95% Confidence Interval was used, which give a sample size result of 362. The number was doubled (approximately) that is, 730 participants in each community as two communities were to be compared in the present study. Individuals of either sex in the age group of 30-65 years were recruited from Kuki (N=730) and Paite (N=730) communities of Churachandpur district, Manipur. Both the studied tribal communities come under Kuki-Chin-Mizo and share numerous similarities-speaks Tibeto-Burman languages, which could be understandable to each other. They originated from Tibet or China via Burma, as per several published records [19]. The geographical location of the Paites is situated in higher altitudes when compared to that of Kuki in the study. Individuals who underwent any major surgery in the recent past, recent blood donation, or any pregnant or lactating women or those suffering from any chronic illness or deformities were excluded from the study.

### Data collection

Data on personal identifiers (name, age, sex) and socio-demography (education, occupation, monthly income) were collected from each participant using interview schedules.

Anthropometric measurements are taken following standard procedures [20].

Height-Standing height was measured using a movable anthropometer. The measurement was done from the vertex (highest point on the head) to the floor.

Weight-It was measured using the digital weighing machine.

Body mass index (BMI) was calculated using weight in kilograms divided by height in meter square.

The concentration of blood hemoglobin (Hb) was measured using an automated AccuSure Hemoglobinometer (HB-101) to measure hemoglobin for anemia status. Capillary blood was collected using a sterile lancet from the participant’s fingertip, the first drop was removed, and the second blood was transferred to the strips set on the device.

### Standard cut-offs

The standardized cut-offs for anemia used are as follows: Males-Normal (≥13 g/dL), Mild (11-12.9 g/dL), Moderate (8.0-10.9 g/dL), severe (<8.0 g/dL). Females-Normal (≥12 g/dL), Mild (11-11.9 g/dL), Moderate (8.0-10.9 g/dL), severe (<8.0 g/dL). In the present study, the number of participants in moderate and severe anemia was low. Hence, these two categories were combined and considered together during analysis [21].

BMI standard cut-offs used as follows: Underweight (<18.5 kg/m^2^), Normal (18.5-22.9 kg/m^2^), Overweight (23.0-24.9 kg/m^2^), and Obese (≥25.0 kg/m^2^) [22].

### Data analysis

All data were analyzed using SPSS version 22. Tests of normality were done for all the studied variables. The median and the Interquartile range (IQR) were used when the data were not normally distributed. Mann-Whitney U test was used to see the difference in median values. Categorical variables were reported as numbers (percentages). The Chi-square test was used to identify differences in categorical variables. The analysis used in this study is frequency distribution to study the prevalence of anemia (total and sex-wise) and among sociodemographic and lifestyle variables. Binary logistic regression was used to estimate the risk of variables for the outcomes. p<0.05 was considered statistically significant.

## Results

The median age of the studied Kuki community was 41.0 (IQR:31.0-54.0) years, whereas, among the Paite community, it was 46.5 (IQR: 34.0-57.0) years. The prevalence of anemia was 42% and 46.4% among Kuki and Paite tribal communities of Manipur, respectively but not statistically significant between the communities. Mild anemia was significantly higher among males in both Kuki (29%) and Paite (35%), whereas moderate/severe anemia was found to be significantly higher among females (21.8% and 22.7%) in both tribal communities. The difference in the prevalence of anemia between males and females in both tribal communities was found to be statistically significant (p<0.001) but not statistically significant when compared between the two communities (Table 1).

**Table 1:**
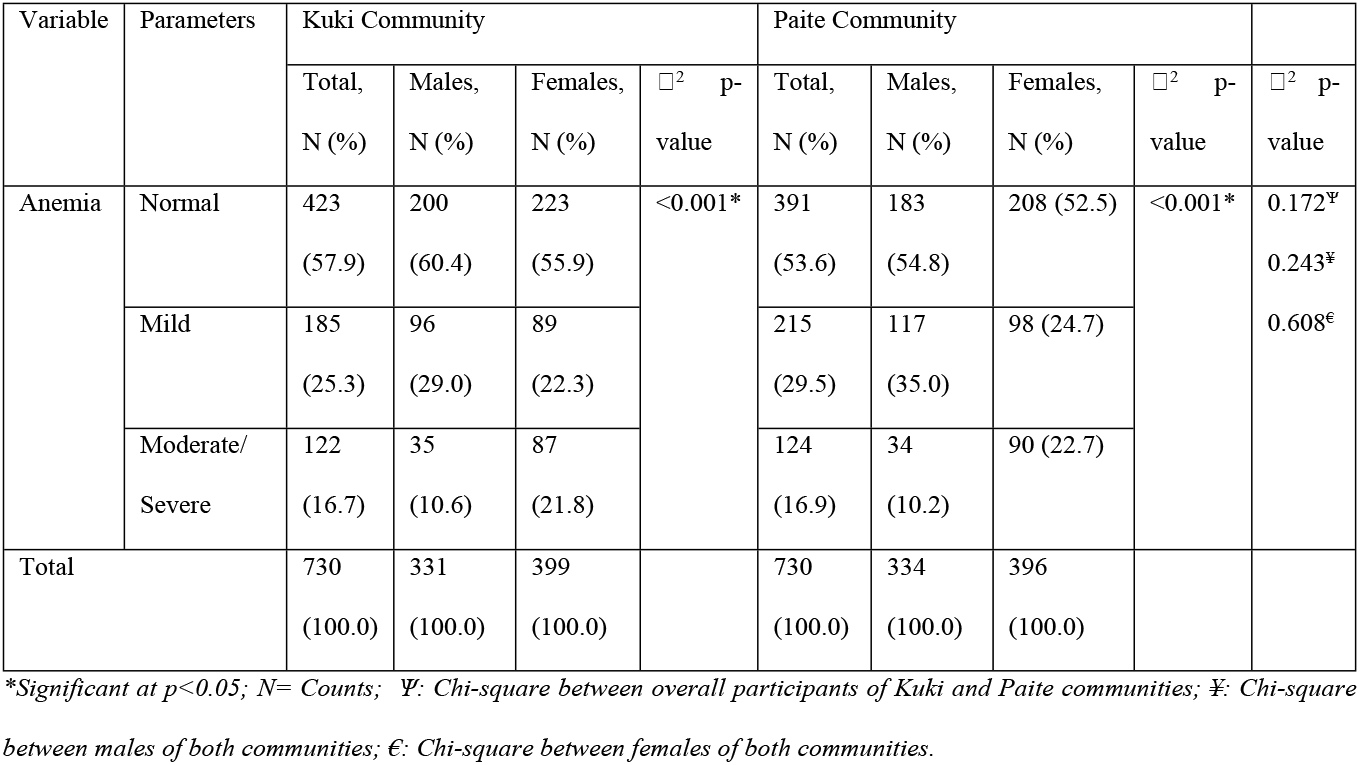
Prevalence of anemia among the two studied tribal communities of Manipur.

The median age concerning anemia was significantly lower in Kuki community than Paite community. In the Kuki community, agriculturist is significantly high among normal and mild anemia, while the homemakers are significantly high in moderate/severe anemia. While in Paite community, agriculturist is significantly high in all categories of anemia. In the studied communities, non-drinkers are significantly high among normal, mild, and moderate/severe anemia (Table 2).

**Table 2:** Distribution of socio-demographic and lifestyle variables with respect to anemia among the two studied tribal communities.

| Variables | Parameters | Kuki Community |  |  |  | Paite Community |  |  |  |
| --- | --- | --- | --- | --- | --- | --- | --- | --- | --- |
|  |  | Normal,<br>N (%) | Mild,<br>N (%) | Moderate/<br>Severe,<br>N (%) | p-value | Normal,<br>N (%) | Mild,<br>N (%) | Moderate/<br>Severe,<br>N (%) | p-value |
| Age | Median (IQR) | 40.0<br>(31.0-51.0) | 43.0<br>(31.5-56.0) | 43 (31.0-60.0) | 0.035* | 46.0<br>(33.0-56.0) | 47.0<br>(35.0-58.0) | 49.0<br>(33.0-60.0) | 0.129 |
| Marital<br>status | Not married | 23 (5.4) | 7<br>(3.8) | 4 (3.3) | 0.559 | 14 (3.6) | 7<br>(3.3) | 3 (2.4) | 0.832 |
|  | Married | 321<br>(75.9) | 144<br>(77.8) | 89 (73.0) |  | 318<br>(81.3) | 170<br>(79.1) | 98 (79.0) |  |
|  | Divorced/<br>widowed (er) | 79<br>(18.7) | 34<br>(18.4) | 29 (23.8) |  | 59<br>(15.1) | 38<br>(17.7) | 23 (18.5) |  |
| Occupation | Unemployed | 56<br>(13.2) | 42<br>(22.7) | 31 (25.4) | <0.001* | 63<br>(16.1) | 47<br>(21.9) | 25 (20.2) | 0.014* |
|  | Agriculture | 142<br>(33.6) | 57<br>(30.8) | 34 (27.9) |  | 177<br>(45.3) | 105<br>(48.8) | 66 (53.2) |  |
|  | Self/government<br>employed | 129<br>(30.5) | 48<br>(25.9) | 19 (15.6) |  | 100<br>(25.6) | 46<br>(21.4) | 15 (12.1) |  |
|  | Home maker | 96<br>(22.7) | 38<br>(20.5) | 38 (31.1) |  | 51<br>(13.0) | 17<br>(7.9) | 18 (14.5) |  |
| Monthly<br>income | ≤10,000 | 271<br>(64.1) | 117<br>(63.2) | 76 (62.3) | 0.194 | 307<br>(78.5) | 174<br>(80.9) | 108 (87.1) | 0.152 |
|  | 10,001-20,000 | 131<br>(31.0) | 61<br>(33.0) | 34 (27.9) |  | 73<br>(18.7) | 34<br>(15.8) | 16 (12.9) |  |
|  | ≥20,001 | 21 (5.0) | 7<br>(3.8) | 12 (9.8) |  | 11 (2.8) | 7<br>(3.3) | 0 (0.0) |  |
| Alcohol | Non drinker | 260<br>(61.5) | 106<br>(57.3) | 92 (75.4) | 0.009* | 269<br>(68.8) | 131<br>(60.9) | 97 (78.2) | 0.001* |
|  | Current drinker | 140<br>(33.1) | 62<br>(33.5) | 26 (21.3) |  | 96<br>(24.6) | 57<br>(26.5) | 14 (11.3) |  |
|  | Ex drinker | 23 (5.4) | 17<br>(9.2) | 4 (3.3) |  | 26 (6.6) | 27<br>(12.6) | 13 (10.5) |  |
| Tobacco consumers | Non consumers | 131<br>(31.0) | 48<br>(25.9) | 34 (27.9) | 0.429 | 86<br>(22.0) | 43<br>(20.0) | 25 (20.2) | 0.815 |
|  | Smoking/Smokeless | 292<br>(69.0) | 137<br>(74.1) | 88 (72.1) |  | 305<br>(78.0) | 172<br>(80.0) | 99 (79.8) |  |
| Smoking status | No | 289<br>(68.3) | 124<br>(67.0) | 90 (73.8) | 0.780 | 169<br>(43.2) | 69<br>(32.1) | 48 (38.7) | 0.053 |
|  | Yes | 131<br>(31.0) | 61<br>(33.0) | 32 (26.2) |  | 206<br>(52.7) | 130<br>(60.5) | 71 (57.3) |  |
|  | Ex-smoker | 3 (0.7) | 0<br>(0.0) | 0 (0.0) |  | 16 (4.1) | 16<br>(7.4) | 5 (4.0) |  |
| Beedis per day | ≤10 beedis | 84<br>(64.1) | 32<br>(52.5) | 22 (68.8) | 0.202 | 137<br>(66.5) | 89<br>(68.5) | 56 (78.9) | 0.145 |
|  | ≥11 beedis | 47<br>(35.9) | 29<br>(47.5) | 10 (31.3) |  | 69<br>(33.5) | 41<br>(31.5) | 15 (21.1) |  |

\*Significant at $p < 0.05$ ; $N = \text{Counts}$ ; Confounders controlled for Kuki communities are age, sex, occupation and alcohol while sex, occupation, alcohol are controlled for Paites.

In Kuki community, underweight participants have a 1.79- and 1.02-folds increased risk of having mild and moderate/severe anemia, respectively, whereas, among Paites, underweight individuals have a 1.65- and 2.19-folds increased risk of having mild and moderate/severe anemia. It is significant only for moderate/severe anemia among the Paites. In the case of overweight/obese individuals, Kuki participants have 0.72- and 0.52-reduced risks of having mild and moderate/severe anemia; while Paite participants have 0.98- and 0.61-reduced risks of having mild and moderate/severe anemia. It is significant for moderate/severe in both communities (Table 3).

**Table 3:** Binary logistic regression of BMI with respect to anemia after controlling for confounders.

| Adiposity indicator | Parameters | Kuki community |  |  |  | Paite community |  |  |  |
| --- | --- | --- | --- | --- | --- | --- | --- | --- | --- |
|  |  | Mild anemia |  | Moderate/Severe anemia |  | Mild anemia |  | Moderate/Severe anemia |  |
|  |  | OR (95% CI) | p-value | OR (95% CI) | p-value | OR (95% CI) | p-value | OR (95% CI) | p-value |
| BMI | Underweight | 1.79 (0.71-4.51) | 0.216 | 1.02 (0.40-2.61) | 0.969 | 1.65 (0.81-3.33) | 0.167 | 2.19 (1.00-4.79) | 0.049* |
|  | Overweight/Obese | 0.72 (0.50-1.06) | 0.093 | 0.52 (0.34-0.81) | 0.004* | 0.98 (0.68-1.42) | 0.918 | 0.61 (0.38-0.98) | 0.039* |

\*Significant at $p < 0.05$ . Confounders controlled for Kuki communities are age, sex, occupation and alcohol while sex, occupation, alcohol are controlled for Paites.

Among the Kuki community, mild anemia trend alongside age tends to show a reverse trend after 50 years. Females in the younger age group (30-39 years) tend to show a high prevalence of anemia which decreases with age. While males, on the other hand, show a high prevalence after 50 years. Whereas among the Paites, females tend to have a high prevalence at an early age, gradually decreasing with age and increasing after 50 years. For males, age is directly proportional to the prevalence of anemia (Fig 1).

**Figure 1:**
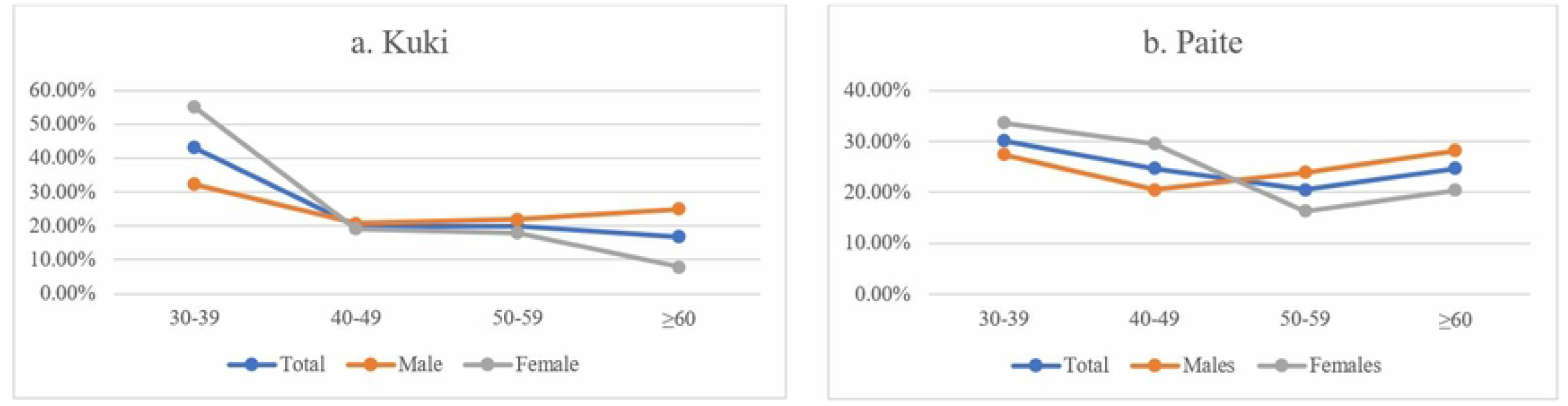
a). Distribution of age cohort (10 years) with mild anemia among Kuki community. b). Distribution of age cohort (10 years) with mild anemia among Paite community.

In the Kuki and Paite communities, the prevalence of moderate/severe anemia is higher among the younger age group (30-39 years), which gradually decreases as the age progresses (40-49 years), after which an increase in the prevalence is seen from 50 years and above. The trend is similar in both males and females. The prevalence of anemia is highest among the female younger age group and the lowest as age increases. However, in the case of males, the prevalence of anemia increased with age (Fig 2).

**Figure 2:**
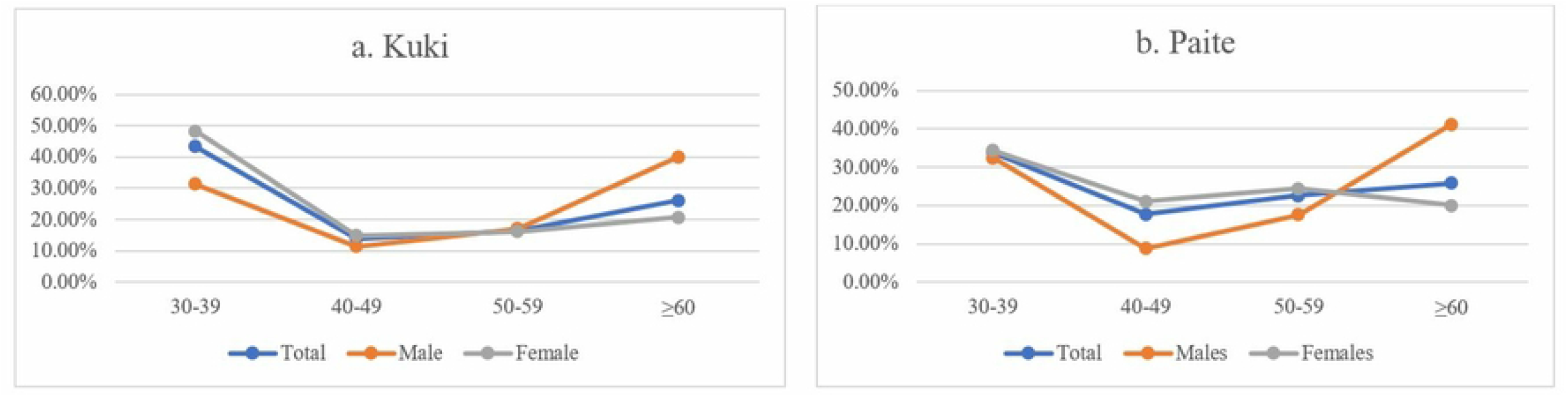
a). Distribution of age cohort (10 years) with moderate/severe anemia among Kuki community. b). Distribution of age cohort (10 years) with moderate/severe anemia among Paite community.

A relationship between underweight, overweight/obese individuals, and anemia was studied with age. It was seen that underweight increases with age while overweight/obese decreases with age in both communities. The prevalence of anemia decreases with age in both the studied communities (Fig 3).

**Figure 3:**
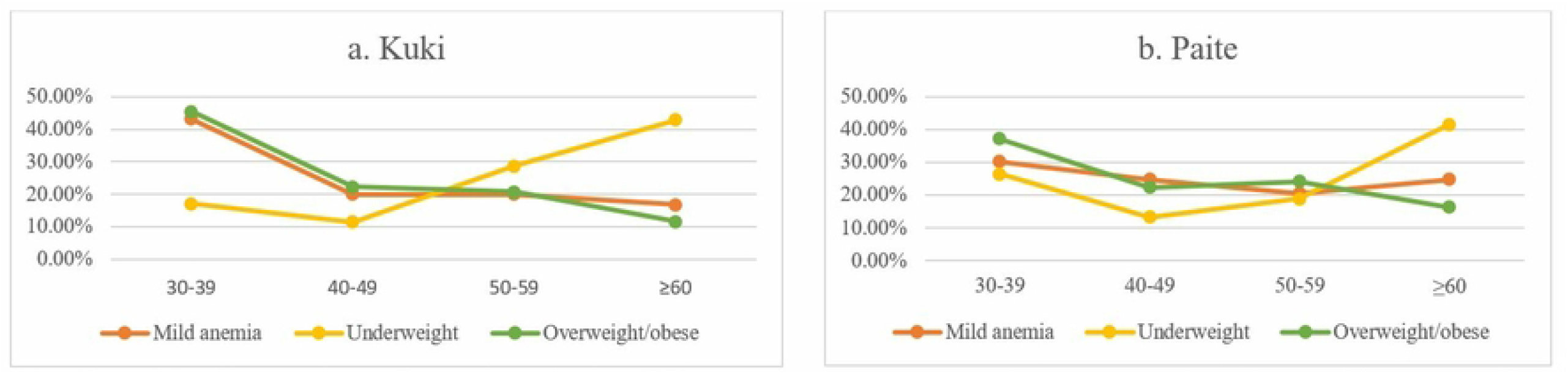
a). Distribution of age cohort (10 years) with mild anemia along with underweight and overweight/obese subjects among Kuki community. b). Distribution of age cohort (10 years) with mild anemia along with underweight and overweight/obese subjects among Paite community.

In the Kuki and Paite communities, the declining trend was observed in overweight/obesity and moderate/severe anemia with age, while underweight increased as age progressed (Fig 4).

**Figure 4:**
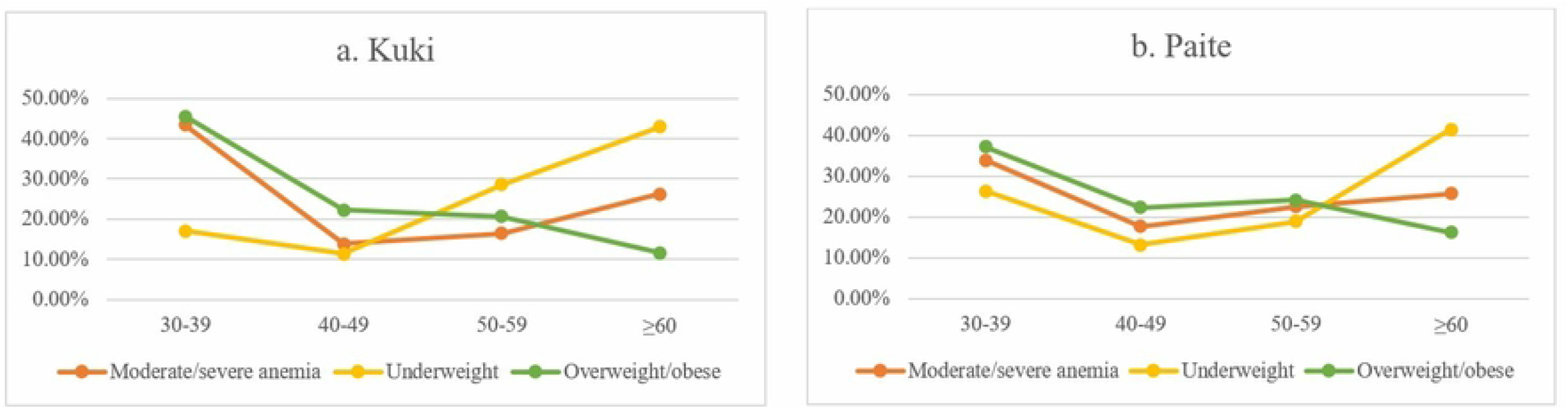
a) Distribution of age cohort (10 years) with moderate/severe anemia along with underweight and overweight/obese subjects among Kuki community. b). Distribution of age cohort (10 years) with moderate/severe anemia along with underweight and overweight/obese subjects among Paite community.

## Discussions

The prevalence of both mild and moderate/severe anemia has been observed for both the tribal communities-Kuki and Paite. The prevalence is higher for Paite community when compared to Kuki community. This might be due to several factors, including nutrition and physical activity [23]. There is an alarming prevalence of underweight among Paites and obesity among Kukis, which need immediate public health interventions. The present study shows that underweight individuals have a significantly higher risk of having anemia (p=0.049) in the case of Paite while among Kukis, there is a high risk but not significant (p=0.069). This result is supported by [24] who reported that underweight individuals are likely to be anemic. The present study shows that being overweight or obese masked the risk of becoming anemic. This is also supported by [25] who reported that an individual who is obese masked off the odds of being anemic.

It was reported that the type of food one consumes also indicates the community [26]. The present study reported a meat consumption disparity between the two tribes, where the frequency of meat consumption is comparatively higher among the Kukis than Paites. Less meat consumption was reported to have a high risk of iron deficiency leading to anemia and vice-versa [27]. Paites have comparatively less meat consumption may be due to the non-availability of meat in markets due to transportation issues, as the studied villages are far away from the heart of the district. The actual scenario is that there is no market; only small shops with limited items can be seen.

Several studies reported that vegetarians, compared to non-vegetarians, have lower hemoglobin counts, which might cause anemia [28, 29]. Also, an essential component of animal food known as Vitamin B-12 plays a vital role in various cellular processes [30] and is abundant in the non-vegetarian diet but lacking in the vegetarian diet. The studies above show that less or no meat consumption, as in the present study case, the Paites, can be more anemic than Kukis.

The present study shows that about 40% of women in both communities are anemic, similar to earlier studies done by [31, 32]. Women were seen to be prioritizing the health of their families first over themselves, which is reported in several studies [33]. Women are an integral part of the family; in most cases, they are solely responsible for preparing food and serving it to families. Women were reported to dine last after their whole family and consume the few leftovers [33]. This might be why females show a high prevalence of anemia due to nutrient deficiency. Also, most women in the present study were found to have high parity, and [5] found that women with high parity have an increased risk for anemia.

Also, anemia shows a declining trend as age progresses in the present study. This might be because women lose heavy blood during the menstrual cycle and childbirth, and have low hemoglobin values during pregnancy [34], thereby causing a high prevalence of anemia [35, 36]. Such anemia arises from iron deficiency, which can be prevented via iron supplements and diets. WHO recommends iron supplements for adolescents and adult women, 30-60 mg of elemental iron to be taken daily for three consecutive months per year [37]. Yet, in the older age group, a natural phenomenon called menopause occurs when women no longer shed blood; hence the prevalence of anemia is low.

This study also shows that Paites have a high rate of illiteracy when compared to Kukis and are more anemic. This is in line with a study done in Ethiopia, stating that education bridged the gap between knowledge, health, and nutrition [38]. The monthly incomes of Paites are comparatively lower than Kukis, and Paites are more anemic in the present study. This study shows a similar trend to other studies [39]. Women’s economic status and anemia [40, 41] were also reported to have an association that supports the present study. A study done among Bangladeshi women found that rural women were more anemic than urban women [36].

It has been reported that smoking and consumption of tobacco products increase the risk of anemia [42–44]. Their results were similar to the present study as it observed high alcohol and tobacco consumption among the studied population. The present study noted that alcohol consumption is higher in Kukis when compared to Paites. The reason comes back to the availability and transportation for the Paites. An additional reason might be because several organizations regularly joined hands to conduct alcohol/tobacco-free-drive in localities to prevent the suppliers and consumers. Smoking of local beedis is comparatively higher among Paites than Kukis. This might be because the Paites are primarily cultivators, and the smoke from beedis acts as mosquito and insect repellent. They are being rolled by the wife or female member of the family for their husband, parents, or siblings, and themselves.

## Conclusions

The high prevalence of anemia, underweight, and overweight/obese individuals in this paper need urgent attention. The present study and existing literature support that anemia is a risk factor for various health complications like cardiovascular diseases (CVDs). Therefore, policymakers must immediately address this issue among the tribal communities. The lack of proper health infrastructure, medical teams, and health policies cause the public to suffer from malnutrition and other diseases. Participants in the present study are tribals - the deprived groups with low literacy rates, high consumers of alcohol and tobacco products, low consumers of nutritious diet and milk, and no proper iron supplementation. The studied tribal communities do not prioritize health much, and several participants never visited a hospital or clinic for a health check-up. Such is the level of ignorance in the communities. To tackle this, more numbers of health institutions with adequate medical staff in rural areas with a minimal charge for a check-up is mandatory and proper functioning should be regularly checked. Proper iron supplementation should be considered as an intervention, as the prevalence of anemia is ≥40% among women in the present study. There should be no/minimal cost for these supplements, and they should reach out to vulnerable populations regularly. Other programs like health awareness programs, literacy programs, and free medical camps should be undertaken. Genetics and biochemical studies should be incorporated into future studies to understand the cause of anemia and suggest appropriate interventions in these rural settings.

## Data Availability

The data will be made available by the corresponding author on request.

## Data availability

The raw data supporting the conclusions of this article will be made available by the authors without undue reservation.

## Ethics statement

The study involving human participants was reviewed and approved by the Ethical Committee of the Department of Anthropology (ref.no.Anth/2022-23/533), University of Delhi.

## Author contribution

JV and MPS designed and executed the study. JV conceptualized the manuscript. JV executed the statistical analyses and wrote the first draft of the manuscript. MPS reviewed and critiqued the statistical analyses and first draft. MPS and JV thoroughly reviewed the final version of the manuscript. All authors contributed to the article and approved the submitted version.

## Acknowledgment

The authors would like to thank all the participants for their time and participation in this study.

